# Ethnicity as a predictor of nonmotor symptoms impact on quality of life in patients with Parkinson’s disease: A systematic review

**DOI:** 10.1101/2023.10.31.23297865

**Authors:** Huda Shujaa Aldeen, Henry Houlden, Mie Rizig

## Abstract

**Background:** Parkinson’s disease (PD) is a chronic progressive hypokinetic movement disorder characterised by rigidity, tremor, and bradykinesia in addition to multiple non-motor features. It is increasingly recognised that NMS have a substantial impact on the quality of life (QoL) of people with PD, more than motor features. The prevalence and types of these NMS vary among PD patients from different ethnic groups. Ethnicity has been postulated as a determinant of PD symptoms and quality of life of people with PD. There is paucity of data on the effect of ethnicity on (QoL) of people with PD.

**Aim:** The aim of this review is to critically evaluate the literature to (1) identify the variability of NMS that affect QoL of people with PD from different ethnicities, and (2) to examine if ethnicity could determine the impact of these NMS on the QoL of people with PD.

**Methods:** This review was conducted according to recommendations in preferred reporting Items for Systematic Review and Meta-Analysis (PRISMA). Search was conducted in three databases (PubMed, Ovid Embase and Scopus) in the period between March & July 2023. NMS were assessed by four scales: NMSS, NMQT, MDS-UDPRS I, and HADS. QoL was assessed by PDQ39. Ethnicity has been defined based on the country where the study has been conducted. A best-evidence synthesis was used to summarise the demographics, study designs, NMS features in each ethnic group. The impact of NMS features on QoL in different ethnic groups was assessed by comparing the power of correlations measures between NMS domains and QoL in all studies.

**Results:** Twenty-one studies with 4246 patients were deemed suitable to be included. Patients in the selected studies were grouped to eight ethnic populations: Caucasian, East Asian, Indian, North African, West African, Hispanic, Mediterranean, and Middle Eastern ethnicities. Heterogeneous protocols and varied statistical methodologies were used in most studies to assess the relation between NMS and QoL of people with PD. Overall depression, fatigue, and sleep and memory problems are the symptoms with the most significant impact on QoL of majority of ethnic populations. Other NMS including cardiovascular, gastrointestinal and urinary Symptoms exhibited a variability in their impact on QoL in different ethnic groups.

**Conclusion:** Our study demonstrates that ethnicity may have a significant impact on the correlation between NMS and QoL of patients with PD. There is deficiency in studies assessing the impact of PD NMS on QoL in Africans, Latin Americans, and Middle Eastern populations. Ethnicity is poorly defined and frequently unaddressed in PD studies. Large multicentre, multinational multi-ethnic studies using rigorous unified protocols are required to properly evaluate the effect of ethnicity on PD NMS. Identification of these variabilities is crucial to aid efficient health policy planning based on each population needs and optimize use of local resources.

## Introduction

Parkinson’s disease (PD) is a chronic progressive neurodegenerative disorder characterised by both motor and non-motor symptoms [1]. PD is the commonest movement disorder, and the second most common neurodegenerative disease after Alzheimer’ s [2]), affecting approximately 6.2 million patients by 2015 [3]). The incidence ofPD increases with age, with around 1% of people over the age of 60 being affected [4]. PD is caused by a combination of genetic and environmental factors [5], with more males being affected than females [6]

Clinically, PD is characterised by cardinal motor features including tremor, bradykinesia (slowness of movement), rigidity and gait disturbance, and postural instability [7]. It was initially considered to be a motor predominant disease; however, in the past two decades, Non-Motor Symptoms (NMS) have also been recognised as predominant features of PD.. A wide range of NMS is now established in PD including: autonomic dysfunction, sleep-related problems, neuropsychiatric symptoms, cognition, pain, and sensory problems. Furthermore, NMS are not only recognised as features of stablished PD, but they may precede PD diagnosis by years and are associated with disease progression and severity.[9]

Motor complications including postural instability and falls [10]; freezing of gait [11]; axial rigidity [12]; dyskinesia [13]; dysphagia (Leow et al., 2010), [15], and motor fluctuations (e.g. wearing-off, morning dyskinesia, biphasic dyskinesia) [16] are widely recognized to be associated with a poor quality of life (QoL) in PD and were considered a major determinant of QoL of people living with PD. However, with the rise in recognition of NMS burden among PD patients, growing number of studies discussed the impact of NMS in PD patients QoL, and deduced that NMS can cause a measurable worsening of the QoL of those individuals during their life course. [17], [18] The NMS include: depression [19], cognitive decline [20], sleep dysfunction [21], excessive daytime somnolence[22], bladder, bowel, and sexual dysfunction [23], and weight loss [24].

PD affects people of all races and ethnicities worldwide [25], [26]. Although there are probably geographic and ethnic differences in the clinical manifestations, epidemiology, and mortality of PD, the exact nature of these differences and their causes remain unclear. Genetic, environmental, and cultural factors [27], [25] could play a role. Understanding of the effect of ethnicity on PD symptoms and their variable effect on QoL of patients with PD is crucial to be able to device appropriate health policies which are tailored to patients’ individual needs. In addition, acknowledging and considering the ethnic diversity in PD is paramount to tackle the global burden of PD worldwide.

Studies assessing ethnic variability in PD manifestations remain very sparse. However, the detection of ethnic variabilities is crucial to targeting the most significant symptoms that affect patients’ daily lives. Recent evidence has implied a close relationship between NMS frequency and severity and HRQoL in PD, with NMS having a greater impact on HRQoL than motor symptoms do [28]. A recent narrative review on ethnic variation in the manifestation of Parkinson’ s disease found that the prevalence of gastrointestinal symptoms appears to be highest in the East Asian studies. The prevalence of depression was above 60% in the Chinese, Korean, Mexican, and Peruvian studies, whereas in the studies from the UK and the USA, the rate was below 40%. However, in a study comparing mood and anxiety symptoms in a multi-ethnic sample, no clear differences were found [29]. Thus, the impact of ethnicity on the presentation of NMS in PD is currently unclear [30]. [31].

This review aimed to systematically examine the effect of NMS on the QoL of PD patients from different ethnic groups. It utilised a systematic methodology to extract and compare studies conducted in varied ethnic populations focusing primarily on those studies measuring the correlation between NMS and QoL of PD patients using standardised and validated clinical scales to identify the NMS domains which are significantly affecting the QoL of PD patients from different ethnic groups, and to compare the variabilities within those domains based on ethnicity.

Most of the current understanding of the determinants of QoL in PD and subsequently the golden standards of PD interventions have been based on data and mat-analyses derived from Caucasian and European populations. According to our knowledge there is no published systematic reviews which assessed the effect of NMS on QoL across different ethnic populations. The findings from this review will give a unique insight to the scale and impact of NMS on QoL in different ethnicities and it will aid tailor the management of patients according to their cultural and individual needs.

## 2. Methods

### 2.1 Protocol

This review was conducted according to recommendations in preferred reporting Items for Systematic Review and Meta-Analysis (PRISMA) [32]. PubMed, Ovid Embase and Scopus were searched in the period between March and July 2023 for literature related to the aim of this systematic review. The review question was formulated using the Population, Intervention, Comparators, Outcome, and Study design (PICOS) framework as follows: **Participants:** PD patients both young onset and old patients. Intervention: None, **Comparison:** Different ethnic groups. **Outcome measure:** Identify the NMS that affect the QoL in each ethnic population. **Comparators**: the group of NMSs which are significantly affecting the QoL of patients from different ethnicities **Study design**: Observational studies; cross-sectional and longitudinal studies that assessed the NMS impact on QoL were included in this review.

PubMed, Ovid Embase, and Scopus were searched for relevant studies. The research question was fragmented into several terms in order to retrieve broad and appropriate results.

The keywords for the three searches were: “Parkinson disease”, “Parkinsons”, “Parkinson’ s disease”, “quality of life”, “questionnaires”, “rating”, “PDQ39”, “NMSS”, “NMSQ”, “HADS”, and “UDPRS”. In PubMed, PD was searched as Medical Subject Headings (MeSH Terms) and text word (tw), and the QoL was searched as MeSh Terms. The same was applied in Ovid where PD was searched as “exp” (exploded) to include MeSh terms. The full search strategy for databases is presented in a*ppendix 1*. Additional relevant articles were added by screening the reference lists of potentially eligible studies.

Literature that satisfied the following criteria was included: Typical idiopathic PD was diagnosed using the UK Brain Bank Criteria UKPDBBC; PDQ39 scale was used to assess the QoL, NMS were measured using NMSS, NMSQ, Hospital and Anxiety Depression score (HADS), MDS-UDPRS Part 1; Studies reporting all NMS domains in each of the scales; Studies should assess both QoL and NMS concurrently in the same population; Each study is conducted in one country, either in one centre or a multicentre, but not in different countries; Papers are written in English or translated into English; Peer-review ed articles; No restriction in time or study design

Exclusion criteria: PD was not diagnosed according to UKPDBBC; Studies not measuring the QoL with PDQ39; Studies assessing NMS by other scales than NMSS, NMSQ, MDS-UDPRS 1, HADS; Studies not reporting on all NMS features in detail; QoL and NMS not assessed concurrently; Papers not written in English or not translated into English; Full article not available; Abstracts, thesis, conference proceedings, posters; Post-mortem studies.

Because most of PD studies do not routinely report on the ethnicity of their studies’ cohorts, names of different ethnicities were not included in the search terms to avoid missing any relevant literatures. Ethnicity was later manually extracted by reading the full text. For this review ethnicity was classified according to the population of the country of study (i.e., we used country of study as proxy for the ethnicity assuming that population of the studies are native to the country of the study unless the author has stated that subjects from different countries were included). Studies not written in English were searched carefully for any available valid translation, and abstracts were searched for their full papers.

Articles were selected by first screening both title and abstract. Subsequently, the potentially eligible articles were read in their entirety and assessed for eligibility. The lists of bibliographies of key articles were also screened to identify any relevant studies. Potentially eligible articles then underwent a quality assessment, as described later, before the article was finally included in the systematic review.

The following data were extracted from the included studies:

- Study design
- Study characteristics (main author, publication year, country of study, recruitment method, Source of participants, sample size)
- Patient demographics (gender, age, age at disease onset)
- Clinical data and assessments used in each study (disease duration, PDQ39, NMSS, NMSQ, H &Y score, MDS UDPRS)
- Mean +SD (or) median IQR of the total score and for each domain of the following scales: NMSS, NMSQ, MDS-UDPRS I.
- PDQ summary index (PDQSI). PDQSI is the sum of dimension total scores of PDQ39 divided by 8.
- Statistical method used by each study to correlate between NMS scales and PDQ39 including:

- Spearman correlation and its *r_s_*
- Pearson’s correlation and the Pearson’ s r
- Multiple regression analysis and the reported [3 coefficient
- Only baseline assessments were extracted from longitudinal studies assessing the NMS progression over several years.

Quality of eligible studies were assessed using a customised validated questionnaire used previously by three systematic reviews assessing the QoL of patients with PD ([33], (Soh and J. L. McGinley et al.(2012) The authors of one of the reviews (Soh and J. L. McGinley et al. (2012) was contacted and consulted on the best approach of using this questionnaires. Briefly, the questionnaire is based on the STROBE checklist [35] and consisted of 14 questions, with each question scoring between zero and two maximum points. The maximum number of points for the questionnaire was 21. A study is considered of high quality if it scores 70% or more of the maximum score.

PDQ39 was used to assess the quality of life in all studies, and measurements were reported as PDQ summary index (SI): this index includes the accumulative scores from eight domains represented by 39 questions. The PDQ SI domains include mobility; activities of daily living; emotional well-being; stigma; social support; cognitive impairment; communication and bodily discomfort.

Non-motor symptoms were assessed by three scales: NMSS in 13 studies, NMSQT in three studies and MDS-UDPRS Part 1 was used in four studies. Both NMSS and NMSQ reported NMS in the following nine domains: cardiovascular symptoms, gastrointestinal dysfunction, urinary symptoms, sexual dysfunction, mood, apathy and cognition, sleep and fatigue, memory and attention problems, hallucinations, and perceptual problems, and miscellaneous (pain, excessive sweating, taste or smell loss, weight loss). NMS were assessed by MDS-UDPRS 1 in 13 domains: cognitive impairment, hallucinations and psychosis, depressed mood, apathy, features of dopamine dysregulation syndrome, sleep problems, daytime sleepiness, pain and other sensations, urinary problems, constipation problems, light-headedness on standing, fatigue.

To compare the differences between NMS and their impact on QoL in PD patients from different ethnicities, a met-regression analysis using individual data of patients was required. However, obtaining this data was not feasible. Therefore, we opted to perform a best evidence synthesis of quantitative and qualitative findings from selected studies. The correlations between NMS domains and QoL in each study were reported including Spearman correlation, Pearson’s correlation, and multiple regression analysis β coefficient, were compared between different studies.

The Spearman correlation coefficient was the most frequently employed statistical method to identify the correlation. The Pearson correlation coefficient was used in three studies, whereas five studies used multiple regression or univariate analysis to predict the impact of NMS domains on QoL. The following were reported in detail in all studies: the r-squared values of the Spearman correlations; the partial r of the Pearson correlations; the Beta coefficient of multiple analysis; and the significant P-value between all NMS domains and QoL.

For the purpose of this review, we crudely defined the ethnicity for each group by the population of the country of study unless the author admitted that he included participants from different countries. In addition, due to the debate over the classification of ethnic groups in medical research, we followed a common classification which roughly identifies the ethnicity by the geographical and social environments. For the ease in describing the different ethnic populations, the term ethnic group will be used frequently.

## 3. Results

The search protocol yielded 9,602 articles. 3397 articles were identified as duplicates by Endnote software and were removed [36].The remaining 6,205 articles were initially screened for their eligibility using the title and abstract: 4,855 articles were excluded following the initial screening. 994 articles were found to be potentially eligible for the inclusion in this review. A meticulous search for full texts was undertaken with 496 had no full article. Full texts were found only for 491 papers. Additional 7 articles were added by screening eligible studies’ reference list. 498 articles were read in full and assessed against the inclusion/exclusion criteria. 477 studies were excluded. Only 21 articles were eligible for inclusion and were assessed qualitatively. *Figure 1: PRISMA flow chart*.

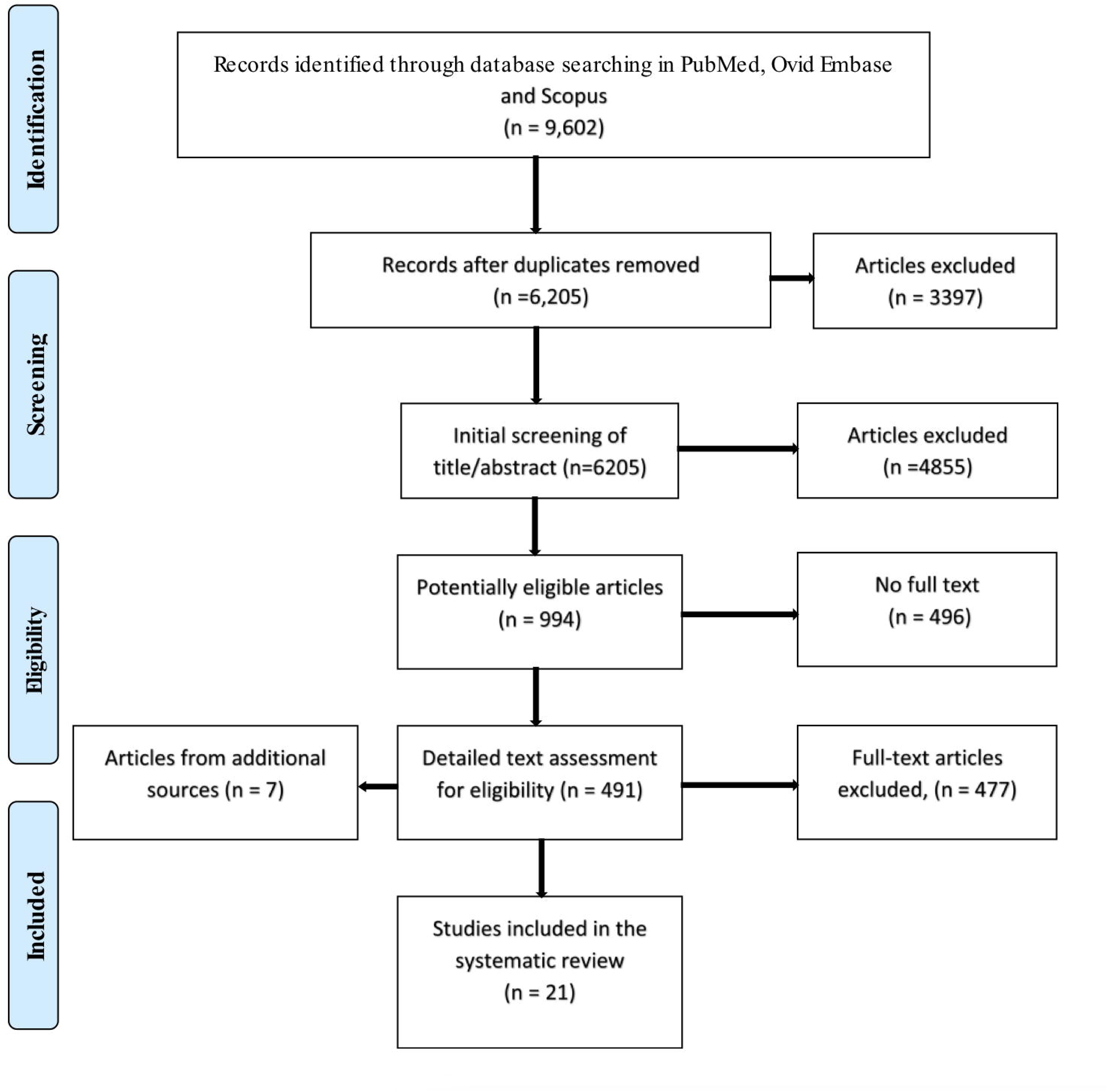

### 3.1 Quality assessment

We have conducted the quality assessment, and all studies were of high quality except one study. Details on the used questionnaire are in *appendix (2)*

### 3.2 Studies characteristics

A total of 21 studies-published between 2010 and 2023 were included in this review. 19 studies were cross-sectional (90%) and two were prospective longitudinal studies (10%). Participants in 19 studies were recruited from a single centre (90%), whereas two studies recruited patients from multiple centres within the same country (10%). Participants in all studies were recruited from hospitals and specialist movement disorder or neurology clinics, except for one study where the patients were recruited from a community-based study[37], [38]

Four studies involved centres in China (Lee *et al.*, 2015) [40] [41] [42] and two in Korea[43] [44] Two studies were conducted in India [45] [46]. In addition, one study was conducted in each of the following countries: United Kingdom (UK) [47], Spain [48], Egypt [49], Nigeria [50]), Singapore [51], Taiwan [52], Malaysia[53], Estonia [37], [38]Slovakia[54]Turkey [55], Iran[56]and Mexico ([57] Studies characteristics were included in ***table 1***.

| Author and Publication year | Country | Ethnicity | Sample size | HRQOL Instrument | NMS Instrument | Correlation method | R2 Score PDQSI/NMS | Statistical analysis |
| --- | --- | --- | --- | --- | --- | --- | --- | --- |
| (Li *et al.*, 2010) | China | East Asian | 82 | PDQ39 | NMSS | Spearman's Rank | 0.67 | Stepwise regression |
| (Song *et al.*, 2014) | China |  | 693 | PDQ39 | NMSS | Spearman's Rank | 0.71 | Logistic regression |
| (Wu *et al.*, 2016) | China |  | 301 | PDQ39 | NMSS | Spearman's/Pearson | 0.61 | Stepwise regression |
| (Hu *et al.*, 2018) | China |  | 569 | PDQ39 | NMSS | Spearman's Rank | 0.62 | Stepwise regression |
| (Kwon *et al.*, 2016) | Korea |  | 323 | PDQ39 | NMSS | Spearman's Rank | 0.33 | Logistic regression |
| (Lee *et al.*, 2015) | Korea |  | 159 | PDQ39 | NMSS | Spearman's Rank | 0.74 | Logistic regression |
| (Liu *et al.*, 2015) | Taiwan |  | 210 | PDQ39 | NMSQ | Spearman's Rank | 0.68 | Stepwise regression |
| (Prakash, 2016) | Singapore |  | 277 | PDQ39 | NMSS |  | NS | Univariate/Multivariate analysis |
| (Azmin *et al.*, 2014) | Malaysia |  | 113 | PDQ39 | NMSS | Spearman's Rank | 0.73 |  |
| (Pappala *et al.*, 2019) | India | South Asian | 64 | PDQ39 | NMSS | Pearson's | 0.71 |  |
| (Karri, Ramasamy and Kalidoss, 2020) |  |  | 100 | PDQ39 | NMSS | Spearman's Rank | - | Multiple regression |
| (Duncan *et al.*, 2014) | UK | Caucasian | 158 | PDQ39 | NMSQ | Spearman's Rank | - | Multiple regression |
| (Liis Kadastik-Eerme *et al.*, 2015) | Estonia |  | 268 | PDQ39 | MDUDPRS | Spearman's Rank | - | Stepwise regression |
| (L Kadastik-Eerme *et al.*, 2016) | Estonia |  | 268 | PDQ39 | MDUDPRS | Spearman's Rank | - | Stepwise regression |
| (Skorvanek, 2015) | Slovakia |  | 291 | PDQ39 | MDUDPRS |  |  | Multiple regression |
| (Berganzo *et al.*, 2016) | Spain | Mediterranean | 103 | PDQ39 | NMSS | Pearson's | 0.56 | Multiple regression |
| (Rodriguez-Violante *et al.*, 2013) | Mexico | Hispanic | 177 | PDQ39 | NMSQ | Spearman's/Pearson's | 0.69 | Multivariate analysis |
| (Gulunay *et al.*, 2020) | Turkey | Middle Eastern | 75 | PDQ39 | NMSQ | Pearson's | NS | - |
| (Salar *et al.*, 2017) | Iran | Middle Eastern | 81 | PDQ39 | NMSS | Spearman's Rank | 0.47 | - |
| (Shalash *et al.*, 2018) | Egypt | North African | 97 | PDQ39 | NMSS | Spearman's Rank | 0.72 | Multiple regression |
| (Arabambi, Oshinaike and Ogun, 2019) | Nigeria | West African | 105 | PDQ39 | NMSS | Spearman's Rank | 0.85 | - |

### 3.3 Patients’ demographics

The total number of patients is 4246 from 15 countries. Most of studies included patients between 100 and 325 (range 103 to 323; median 193.5). 5 studies included less than 100 patients (range 64-97), while two studies included over 500 participants. The included patients are covering eight ethnicities, Caucasian, East Asian, South Asian, North African, West African, Hispanic, Mediterranean, and Middle Eastern. *Figure 2*: *Studies locations and ethnic populations*. East Asian ethnicity included studies from China, Korea, Taiwan, Singapore, and Malaysia. Caucasians were represented by studies from three countries, the UK, Slovakia, and Estonia. Mediterranean people were represented by a study from Spain. African studies were divided into North African from Egypt, and West African from Nigeria, and a study from Mexico was included as Hispanic ethnicity. Both Iranian and Turkish studies were grouped as Middle Eastern. Furthermore, one study from India was included as South Asian. We did not find people from South Africa, America and Arab.

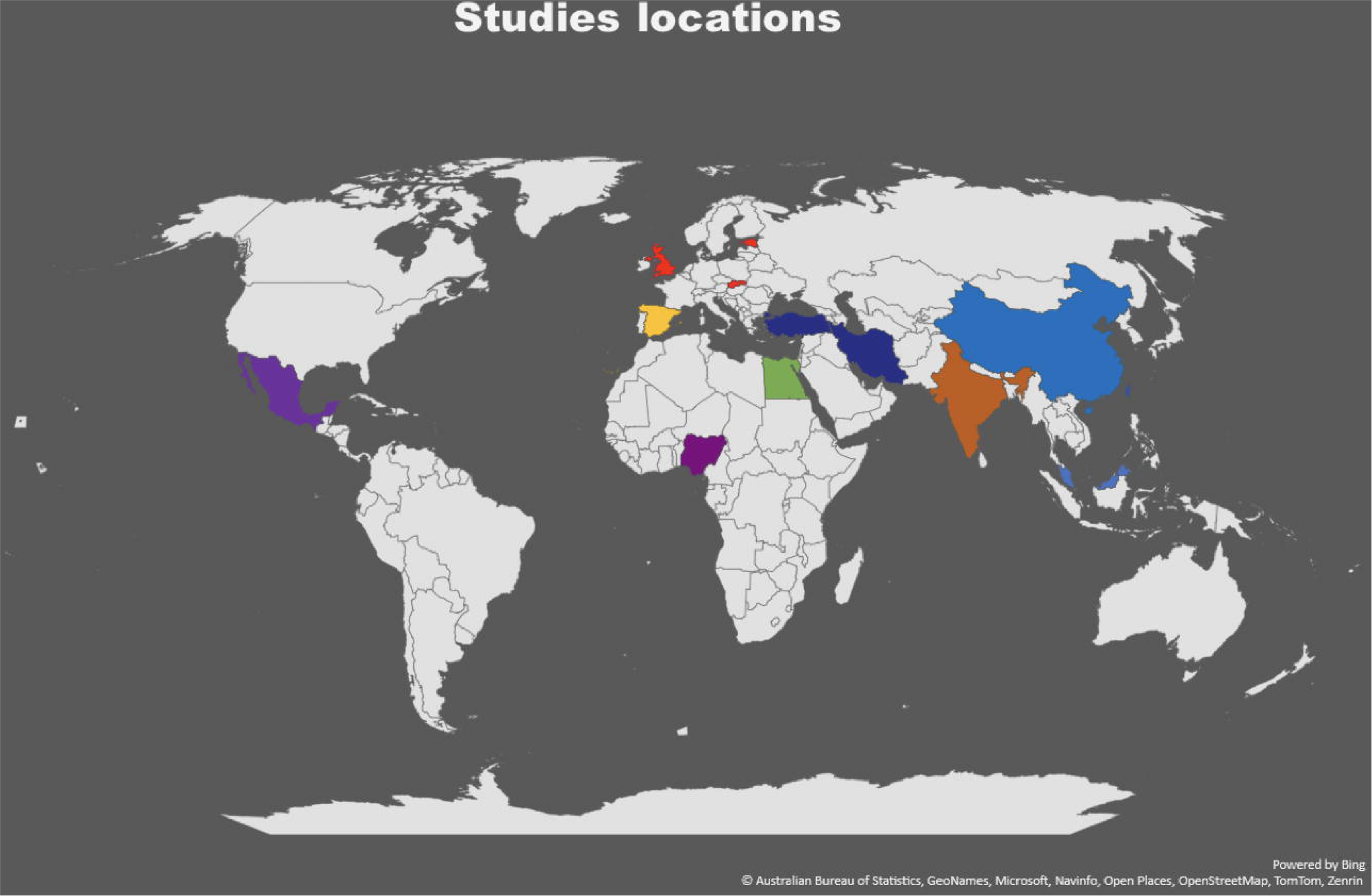

The ratio of males to females is high in most of studies 1.1/0.9 and the main age of patients is 64.35 years with the majority of them above 55 years, so most of included patients are old-onset PD. Furthermore, a study from Estonia included older patients with a mean SD age of (74.2 ±8.8) [37], [38]Twelve studies reported the mean age at disease onset which was 58.36 years (range 50.1 to 66.8). The mean disease duration in most studies was 5.93, and a range of means (4.1 to 8.3) except in four studies where the mean disease duration ranged from 0.5 to 3 years[47], (Lee *et al.*, 2015) [40] [42]. The shortest disease duration was in s study from the UK which included newly-diagnosed patients]. while the longest disease duration was in studies from Slovakia and Estonia.

The motor assessment with UDPRS III showed that Estonian patients had the worst score because they were elder than the patients in other studies and with longer disease duration which points that patients with a long disease duration may experience more motor impairment. Most of the patients were on treatment except in two studies which included drug naïve PD patients (Lee *et al.,* 2015), [42] Patients’ demographics ***table 2***

| Author and Publication year | Country | Ethnicity | Study design | Recruitment method | Source of participants | Diagnostic Criteria | Sample size | M:F | Mean Age,yr (rang) | Mean age at oncet | Mean disease duration Year | Mean HY stage (range) | UDPRS III (Mean±SD) Median (IQR) | HRQoL Scale | NMS scale | Correlation method | Statistical analysis |
| --- | --- | --- | --- | --- | --- | --- | --- | --- | --- | --- | --- | --- | --- | --- | --- | --- | --- |
| (Li *et al.*, 2010) | China | East Asian | CS | NS | Neurology department | UKPDSBBC | 82 | 46/36 | 65 | NS | 5.1 | 2* | 16.9 ±7.0 | PDQ39 | NMSS | Spearman's Rank | Stepwise regression |
| (Song *et al.*, 2014) | China |  | CS | Consecutive | Neurology clinic/Neurology department | UKPDSBBC | 693 | 386/307 | 61.5 | 57.1 | 4.4 | 2.5 | 30.2 ± 13.4 | PDQ39 | NMSS | Spearman's Rank | Logistic regression |
| (Wu *et al.*, 2016) | China |  | CS | NS | Neurology department | UKPDSBBC | 301 | 165/136 | 58.4 | 56.3 | 2.1 | 1.9 (I-II) | 22.3 ± 10.3 | PDQ39 | NMSS | Spearman's/Pearson | Stepwise regression |
| (Hu *et al.*, 2018) | China |  | CS | NS | Neurology department | UKPDSBBC | 569 | 275/294 | 58.1 | 56.1 | 2 | 1.9 | 21.7 ± 11.8 | PDQ39 | NMSS | Spearman's Rank | Stepwise regression |
| (Kwon *et al.*, 2016) | Korea |  | CS | Consecutive | Movement disorder clinics | UKPDSBBC | 323 | 153/170 | 66.8 | 62.8 | 4.1 | 2.4 | 24.9±14.2 | PDQ39 | NMSS | Spearman's Rank | Logistic regression |
| (Lee *et al.*, 2015 | Korea |  | CS | NS | Movement disorder clinic | UKPDSBBC | 159 | 66/93 | 68 | NS | 1.4 | 2.2 (I-V) | 22.6±10.5 | PDQ39 | NMSS | Spearman's Rank | Logistic regression |
| (Liu *et al.*, 2015) | Taiwan |  | CS | NS | Movement disorder clinic | UKPDSBBC | 210 | 110/100 | 66.1 | 60 | 6.1 | 2.2 (I-V) | NS | PDQ39 | NMSQ | Spearman's Rank | Stepwise regression |
| (Prakash, 2016) | Singapore |  | Longitudinal | NS | Movement disorder clinics | UKPDSBBC | 277 | 136/141 | 64.4 | NS | 5.8 | (I-V) | 27.0 (11.0, 36.0) | PDQ39 | NMSS |  | Univariate/Multivariate analysis |
| (Azmin *et al.*, 2014) | Malaysia |  | CS | NS | Neurology clinic | UKPDSBBC | 113 | 60/53 | 64.8 | 59 | 5 (2–8) | 2* (I-IV) | NS | PDQ39 | NMSS | Spearman's Rank |  |
| (Pappala *et al.*, 2019) | India | South Asian | CS | NS | Neurology clinic | UKPDSBBC | 64 | 43/21 | 56.1 | NS | 3.1 | 2.2 (I-V) | NS | PDQ39 | NMSS | Pearson's |  |
| (Karri, Ramasamy and Kalidoss, 2020) |  |  | CS | NS | OPD/Inpatient | UKPDSBBC | 100 | 66/34 | 68.4 | NS | 3.5 | 2.95 | NS | PDQ39 | NMSS | Spearman's Rank |  |
| (Duncan *et al.*, 2014) | UK | Caucasian | Longitudinal | NS | Movement disorder, neurology, and medicine OPD clinics | UKPDSBBC | 158 | 104/54 | 66.5 | NS | 0.5 | I-IV | 27.1±12 | PDQ39 | NMSQ | Spearman's Rank | Multiple regression |
| (Liis Kadastik-Eerme *et al.*, 2015) | Estonia |  | CS | Community | Neurology department | UKPDSBBC | 268 | 105/163 | 74.2 | 66.8 | 7.6 | 3* (I-V) | 46.3±18.3 | PDQ39 | MDS-UDPRS I | Spearman's Rank | Stepwise regression |
| (L Kadastik-Eerme *et al.*, 2016) | Estonia |  | CS | Community | Neurology department | UKPDSBBC | 268 | 105/163 | 74.2 | 66.8 | 7.6 | 3* (I-V) | 46.3±18.3 | PDQ39 | MDS-UDPRS I | Spearman's Rank | Stepwise regression |
| (Skorvanek, 2015) | Slovakia |  | CS | NS | Movement disorder centre | UKPDSBBC | 291 | 156/135 | 68 | 59 | 8.3 | 2.7 (I-V) | 39.1±16.6 | PDQ39 | MDS-UDPRS I |  | Multiple regression |
| (Berganzo *et al.*, 2016) | Spain | Mediterranean | CS | Consecutive | Movement disorders clinic | UKPDSBBC | 103 | 55/48 | 66.5 | NS | 6.9 | 2.3 | 30.0±9.9 | PDQ39 | NMSS | Pearson's | Multiple regression |
| (Rodriguez-Violante *et al.*, 2013) | Mexico | Hispanic | CS | NS | Movement disorder clinic | UKPDSBBC | 177 | 92/85 | 62.4 | 56 | 6.4 | (I-V) | 23.3±13.1 | PDQ39 | NMSQ | Spearman's/Pearson | Multivariate analysis |
| (Gulunay *et al.*, 2020) | Turkey | Middle eastern | CS | Consecutive | Neurology clinic | UKPDSBBC | 75 | 65/29 | NS | 58.8 | 5.1 | 2.1 | 13.6±8.4 | PDQ39 | NMSQ | Pearson's |  |
| (Salar *et al.*, 2017) | Iran | Middle eastern | CS | NS | PD clinic | UKPDSBBC | 81 | 60/21 | 62 | NS | 6.1 | (I-V) | 35.0 ± 18.0 | PDQ39 | NMSS | Spearman's Rank |  |
| (Shalash *et al.*, 2018) | Egypt | North African | CS | NS | Movement disorder clinic | UKPDSBBC | 97 | 58/39 | 55.3 | 50.1 | 5.3 | 2.8 (I-V) | 38.04±19.4 | PDQ39 | NMSS | Spearman's Rank | Multiple regression |
| (Arabambi, Oshinaike and Ogun, 2019) | Nigeria | West African | CS | NS | Neurology clinic | UKPDSBBC | 105 | 65/40 | 64 (55–70) | NS | NS | (I-V) | NS | PDQ39 | NMSS | Spearman's Rank |  |

### 3.3 Clinical assessments

#### 3.3.1 Disease Progression (HY stage)

The mean Hoehn and Yahr (H&Y) stage was 2.29, and most of the patients in all included studies were in the mild to moderate stage HY (I_III). The studies also included patients in the advanced stage (IV, V) who represented less than 10% of the study population, except in two studies; North African with advanced PD representing 21% [49] and East Asian with HY stage at three or more, representing 28% [44]. Most studies did not report whether the patients were assessed in the on or off status except for two studies, one of which evaluated the patients in both the on and off status [49] and the other assessed the patients in the on status only [47]. Motor symptoms were assessed with UDPRS III in 16 studies, whereas 4 studies did not assess the motor symptoms of the patients.

#### 3.3.2 Burden of PD NMS in different ethnic groups

NMSS scores were compared only in 9 studies which had comparable method of measurement mean SD. The ethnicities being compared in this section are Middle Eastern, East Asian, South Asian, Mediterranean, and North African. Other ethnicities and some studies were not compared here for reasons discussed later. The North African group reported the highest NMS score of 61.0+42.9, whereas Mediterranean patients had a high score of 42.7±24.7. The Middle Eastern and East Asian groups reported lower scores of 37.0+22.5 and 35.3+33.3, respectively. In South Asian group, one study has reported the highest NMSS total score 72.8+44.2, while the other study showed the lowest NMSS score 30.8± 29.1 among all studies populations and ethnicities. In East Asian countries, one Korean study reported the highest NMSS score among other Korean and Chinese studies[44], as they included a high number of patients in the advanced stage, representing 28% of the total number of study population. ***Table 3*** illustrates the studies for a comparison of NMS scores between different ethnicities.

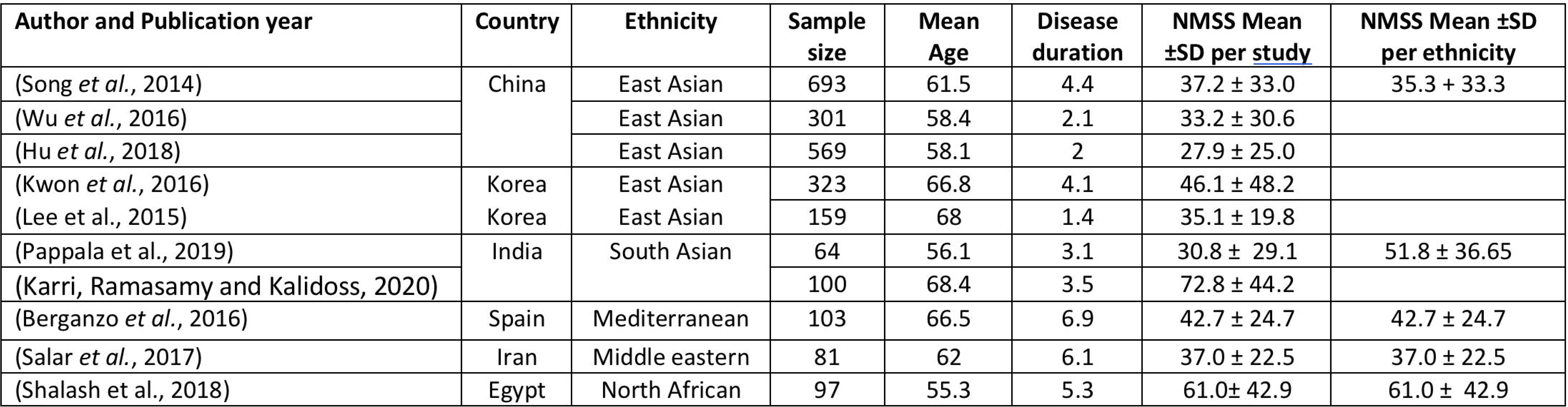

#### 3.3.4 QoL in patients with PD from different ethnic groups

We have decided to report the PD-QSI scores in order to compare the QoL of PD patients between different ethnicities, as it was reported in most of studies to summarize the total PDQ39 scoring. Studies from six ethnicities had reported the PDQSI were compared for the QoL variation in PD patients. The worst QoL score was of the Middle Eastern and North African and the best was of the East Asian. Middle Eastern and North African patients exhibited the worst QoL among all ethnicities with a PDQSI of 37.5 ±18.6 and 38.2+29.3 respectively. This was followed by the Hispanic and Caucasian groups, scoring 33.52 ±19.04 and 29.51 ± 18.51, respectively. The East Asian and Mediterranean groups had the lowest PDQSI scores; thus, they have better QoL scores. ***Table 4*** Although the South Asian study was not included in the ethnicity comparison of the QoL as it did not report the PDQSI score, but they reported a high total PDQ39 score of 158.3 ± 114.07. [45] It is worthy of mention that only one Chinese study reported a very high PDQSI score of 43.0±14.7, because of the high number of patients in advanced PD in comparison to the other studies in this review. (Lee *et al.*, 2015)

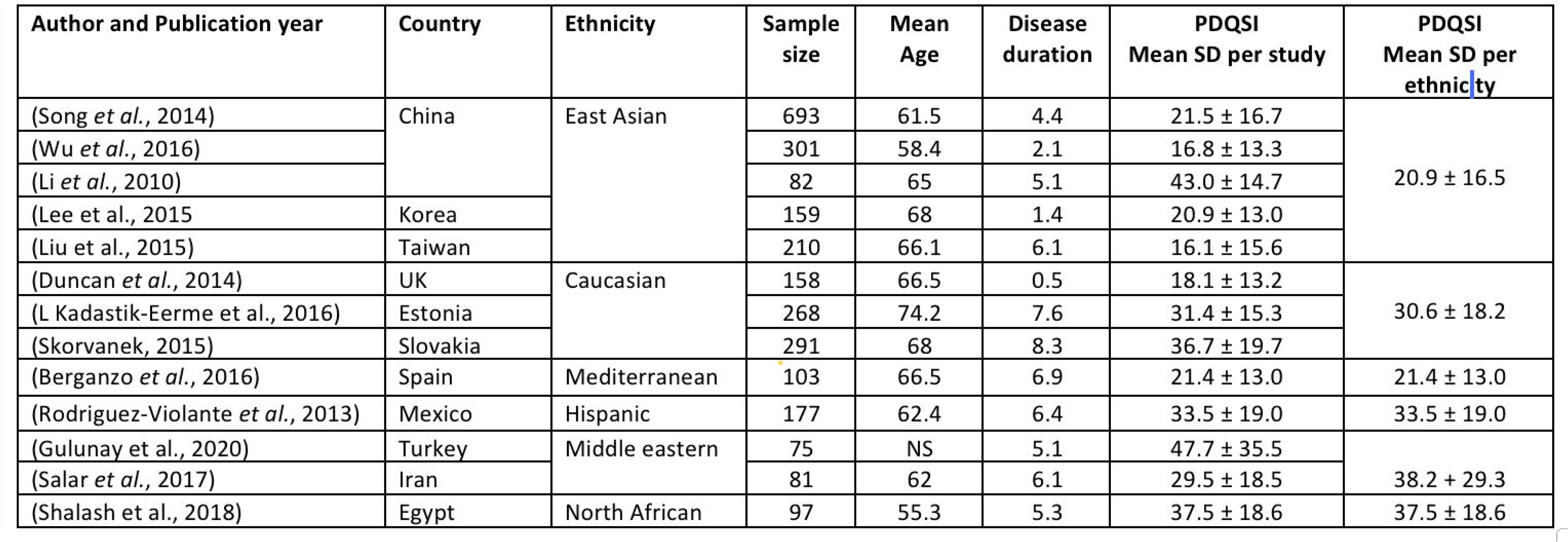

| Author and Publication year | Country | Ethnicity | Sample size | Mean Age | Disease duration | PDQSI Mean SD per study | PDQSI Mean SD per ethnicity |
| --- | --- | --- | --- | --- | --- | --- | --- |
| (Song *et al.*, 2014) | China | East Asian | 693 | 61.5 | 4.4 | 21.5 ± 16.7 | 20.9 ± 16.5 |
| (Wu *et al.*, 2016) |  |  | 301 | 58.4 | 2.1 | 16.8 ± 13.3 |  |
| (Li *et al.*, 2010) |  |  | 82 | 65 | 5.1 | 43.0 ± 14.7 |  |
| (Lee *et al.*, 2015) | Korea |  | 159 | 68 | 1.4 | 20.9 ± 13.0 |  |
| (Liu *et al.*, 2015) | Taiwan |  | 210 | 66.1 | 6.1 | 16.1 ± 15.6 |  |
| (Duncan *et al.*, 2014) | UK | Caucasian | 158 | 66.5 | 0.5 | 18.1 ± 13.2 | 30.6 ± 18.2 |
| (L Kadastik-Eerme *et al.*, 2016) | Estonia |  | 268 | 74.2 | 7.6 | 31.4 ± 15.3 |  |
| (Skorvanek, 2015) | Slovakia |  | 291 | 68 | 8.3 | 36.7 ± 19.7 |  |
| (Berganzo *et al.*, 2016) | Spain | Mediterranean | 103 | 66.5 | 6.9 | 21.4 ± 13.0 | 21.4 ± 13.0 |
| (Rodriguez-Violante *et al.*, 2013) | Mexico | Hispanic | 177 | 62.4 | 6.4 | 33.5 ± 19.0 | 33.5 ± 19.0 |
| (Gulunay *et al.*, 2020) | Turkey | Middle eastern | 75 | NS | 5.1 | 47.7 ± 35.5 | 38.2 + 29.3 |
| (Salar *et al.*, 2017) | Iran |  | 81 | 62 | 6.1 | 29.5 ± 18.5 |  |
| (Shalash *et al.*, 2018) | Egypt | North African | 97 | 55.3 | 5.3 | 37.5 ± 18.6 | 37.5 ± 18.6 |

All scales assessing the NMS were reported, and the correlation between the QoL and NMS analysed by different analysis and correlation methods was reported. The total NMS scores measured by different scales were significantly correlated with QoL in all ethnicities.

##### A) East Asian group

Mood/cognition and sleep/fatigue symptoms were significantly correlated with QoL in all East Asian studies from China, Korea, Taiwan, Malaysia and Singapore. (Lee *et al.*, 2015) [40] [41] [42] [43] [44] [52] [51] [53] However, the strength of the correlation varied among studies, with the strongest significant correlation for sleep being reported by [41], other detected significant moderate correlation. By multivariate analysis, sleep/fatigue was correlated at P 0.0008 in a study from Singapore. [51]

Attention/Memory showed variability among East Asian studies, with only two studies reported a strong significant correlation with the QoL (Lee *et al.*, 2015), [51]. Also, a moderate significant correlation was detected by [40] [41] [44] [52]

The remaining studies identified weak or non-significant correlations. Li *et al.*, 2010), [40], [53] Gastrointestinal symptoms were correlated significantly with the QoL, but with a moderate correlation coefficient in a study from each China and Korea [41], [44]. All other East Asian groups reported a significant but weak correlation between GIT symptoms and QoL. Furthermore, only one study highlighted a significant moderate correlation between urinary symptoms and QoL (Lee *et al.*, 2015). The same study reported a strong significant correlation between the miscellaneous symptoms and the QoL. Another study has reported a significant moderate correlation with the QoL [41]. While all other Asian studies reported either a non-significant or weak correlation with these two NMS domains. Cardiovascular symptoms were signifyingly correlated with PDQ39 in a study from Malaysia, and these symptoms were reported as the major NMS affecting the QoL in this study [53] In addition, significant moderate correlation between the cardiovascular symptoms and the QoL was also reported in other two studies. [44][52]. ***Table 5*** demonstrates the most significantly correlated NMS in each ethnicity.

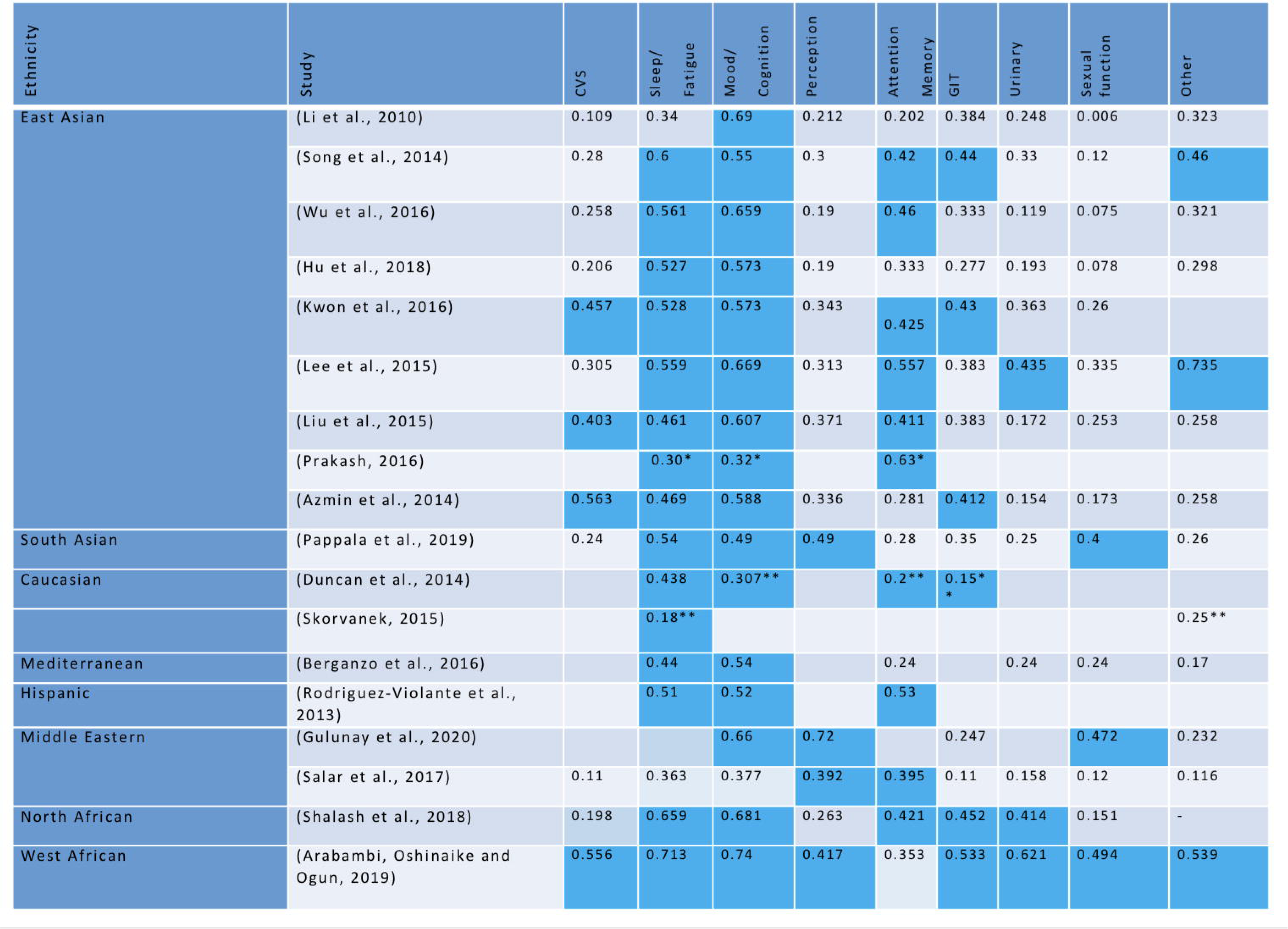

##### B) South Asian (Indian) group

By the Pearson correlation coefficient, all NMS showed a significant correlation with the quality of life with P-value <0.05; however, the strongest correlations were between the QoL and sleep and fatigue, followed by mood and cognition, hallucination, and perceptual problems with *P* =0.00, rs =0.54 and *r_s_*=0.49, receptively. Furthermore, the sexual dysfunction and GIT symptoms domains correlated at *P*=0.00 with *r_s_* =0.4 and *r_s_*=0.35, respectively.[45]. By Spearman’s correlation, another Indian study reported strong significant correlation between QoL and mood, attention and gastrointestinal symptoms domains with *P*= 0.00 in 100 patients, in addition, cardiovascular symptoms and sleep dysfunction showed significant correlation with QoL with P= 0.001 and 0.002 respectively. However, the perceptual, urinary and sexual domains had less or non-significant impact on the QoL with *P*= 0.006, 0.536 and 0.658 respectively. [46]

##### C) The Caucasian group

A study from the UK analysed the connection between the QoL and NMSQ domains by the stepwise multiple linear regression and the Spearman correlation. This study found that the NMS that significantly affected the QoL in PD were depression ([3=0.307); impaired concentration ([3=0.204; P<0.001); anxiety ([3 =0.156); and the sensation of incomplete bowel emptying ([3=0.153; P=0.014). Whereas by Spearman correlation, attention/memory problems and sleep disturbance showed the strongest significant correlation with QoL in addition to Depression and Anxiety. Furthermore, NMSQ items of “low mood,” “daytime sleepiness,” and “insomnia” were rated with the GDS-15, ESS, and PSQI, and showed significant correlations with: GDS-15 (Pearson’ s r=0.611; *P*<0.00; PSQI (Pearson’s r=50.438; *P*<0.001); and ESS scores (Pearson’ s r=0.373; *P*=0.001). [47] Both Slovakian and Estonian studies assessed NMS by using MDS-UDPRSI. In the Slovakian study which used multiple linear regression, QoL was significantly affected by pain ([3=0.25, *P* = <0.0001), followed by fatigue ([3=0.18, *P* = <0.001) and features of dopamine dysregulation syndrome (DDS), ([3=0.10, *P*= <0.01). [54]In the Estonian study, which used Spearman correlation, all NMS features, except ICD, were correlated significantly with QoL. However, the r-squared of these correlations was not reported. Moreover, depression was assessed separately by BDI and showed a strong significant correlation with PDQSI Spearman’s correlation coefficient (CC=0621 *P*=<0.0001). Depression was also reported as the most significant determinant of QoL in this study. [37], [38]

##### D) The Mediterranean group

By the Pearson correlation coefficient, the QoL correlated significantly with only two domains, mood and cognition, and sleep, and fatigue, with (*P*<0.001) and CC=0.54 and CC=0.44, respectively. All other domains exhibited non-significant correlations. [48]

##### E) The Hispanic group

By multiple linear regression with the standardized beta being reported, the main non-motor domains affecting the quality of life were depression/anxiety ([3 = 6.36), cardiovascular ([3 = 5.39), memory ([3=4.64), and miscellaneous ([3 = 3.15). Correlations between NMS domains and QoL were also analysed by the Spearman technique. The strongest significant correlations were between the QoL and apathy/attention/memory (*r_s_*=0.53), depression/anxiety (*r_s_* = 0.52) and sleep disturbances (*r_s_*=0.51). [57]

##### F) The Middle Eastern group

By the Pearson correlation coefficient, the Turkish study reported the strongest significant correlations between the QoL and the sexual dysfunction among patients under 60 years of age (Pearson’s *r* =0.67), and perception disorder among patients over 60 (Pearson’ s *r* =0.72). Furthermore, depression and sleep were assessed separately by GDS and PDSS, and a strong significant correlation was found between depression and QoL (Pearson’s r=0.66, *P*< 0.01), while sleep had a moderate correlation with QoL PDSS (Pearson’s r=0.38). [55], In addition, by Spearman’s rank correlation coefficient, a study from Iran showed that the strongest two domains which correlated significantly with the QoL were Attention/Memory and Perceptual problem/hallucination at *P* <0.001 and *r_s_* =0.395, *r_s_*=0.392, respectively. However, mood and cognition, sleep, and fatigue correlated with QoL at *P* =0.001 and *r_s_*=0.377, *r_s_*=0.363, respectively. [56]

##### G) The North African group

By the Spearman correlation coefficient, the strongest two domains that correlated significantly with QoL were mood/cognition, followed by the sleep/fatigue domain with *r_s_*=0.681 and *r_s_*=0.659, respectively. Furthermore, digestive symptoms, attention and memory, and urinary symptoms exhibited a moderate significant correlation with *r_s_*=0.452, *r_s_*=0.421 and *r_s_*=0.414, respectively. All other domains showed non-significant correlations. [49]

##### H) The West African group

By Spearman correlation coefficient, all NMS features were correlated significantly with QoL. However, mood/cognition, and sleep/fatigue had the strongest correlation with *r_s_*=0.740 and *r_s_* =0.713, respectively. All other features displayed moderate significant correlation except for hallucination and perceptual problems domain which had weak correlations with QoL. The moderate significant correlation were reported in urinary *r_s_*=0.621, cardiovascular *r_s_*=0.556, miscellaneous *r_s_* =0.539, gastrointestinal *r_s_*=0.533, perceptual problems/hallucinations *r_s_*=0.417, sexual function *r_s_*=0.494, and attention/memory *r_s_*=0.353. [50]

#### 3.3.3 Effect of ethnicity on NMS by domain

##### 1. Cardiovascular symptoms

CVS was correlated significantly with QoL in the Hispanic and West African studies, with three East Asian studies from Malaysia, Korea and Taiwan [44][52] [53] CVS were reported as the major NMS impacting the QoL in both the Hispanic and Malaysian studies. [57], [53] There was no significant correlation between CVS and QoL in all other ethnic categories.

##### 2. Sleep and fatigue

Sleep and fatigue problems were strongly correlated with QoL in all evaluated ethnicities in this review: East and South Asian, Caucasian, Mediterranean, Hispanic, and both North and West African. Furthermore, fatigue was assessed separately by MDS-UDPRS I and exhibited a significant correlation with QoL in Slovakian patients. [54], No significant correlation was observed between sleep/fatigue and QoL in Middle Eastern patients from Turkey by using Spearman correlation between NMSQ and PDQ39. However, sleep was assessed separately by a different scale (PDSS) and a significant but weak correlation was identified. [55] In addition, Iranian patients showed a non-significant connection between QoL and sleep and fatigue domain. [56]

##### 3. Mood and cognition

Mood, in particular, depression, was significantly correlated with QoL in almost all ethnicities included in this review. In the Middle Eastern group, depression was assessed separately by GDS in Turkish patients, and they found a significant impact of depression on QoL. [55] However, in the Iranian study, a significant but moderate to weak correlation *r_s_*=0.39 was reported and the author has not commented on the correlation results. [56] No significant correlation was reported in the Slovakian population, depression as measured by MDS-UDPRS I exhibited a non-significant correlation with QoL measured by PDSI score in the non-fluctuating patients’ group. However; depression showed a strong correlation with QoL in the fluctuating group. [54]

##### 4. Perceptual problems and hallucinations

The Middle Eastern, South Asian, and West African groups showed a significant positive correlation between hallucinations and perceptual problems domain and QoL. [55], [56]**_**[45] [50]

##### 5. Attention and memory

QoL was significantly affected by impaired attention and memory problems in most of the ethnicities. The Asian, Caucasian, North African and Middle Eastern, Iranian only, groups displayed significant correlations. By multiple linear regression, attention and memory problems had a significant impact on QoL in Hispanic patients. [57] Nevertheless, no or non-significant correlations were observed in the Mediterranean and Turkish patients, whereas the South Asian and West African groups exhibited a significant, but weak, correlation.

##### 6. Gastrointestinal tract

GIT symptoms were correlated significantly with QoL, but with moderate *r_s_* in many studies from the East Asian group and one south Asian study. [43] [44], [46] [53] In addition, two studies from the Caucasian group, the UK and Estonia reported significant correlation while in the Slovakian study non-significant correlation was identified. [37], [38] [54]The North African group displayed a significant moderate correlation, although this was not reported by authors. All other ethnic groups showed either a weak or non-significant correlation. [49]

##### 7. Urinary Symptoms

Urinary symptoms were not reported as a determinant of QoL in any of the reviewed ethnicities. However, by spearman correlation the North, West African and one Chinese study reported a significant moderate, correlation between the urinary symptoms and the quality of life[50], [49], (Lee *et al.*, 2015)

##### 8. Sexual dysfunction

Middle Eastern people, particularly Turks, were the only ethnic category in which sexual dysfunction was reported to have had a significant impact on the QoL, [55] whereas no significant correlation was reported in Iranian patients. Furthermore, the West African study reported a significant moderate correlation between the sexual dysfunction domain and QoL; however, this symptom was not reported as a major determinant of QoL among the NMS.[50]

##### 9. Miscellaneous

The miscellaneous domain includes four main symptoms: weight loss, pain, loss of taste or smell, and excessive swelling. The correlation between QoL and the miscellaneous domain was significant only in the West African and East Asian, two studies only[50], (Lee *et al.*, 2015), [41]Pain and DDS assessed separately in Slovakian patients with MDS-UDPRS I, showed a very significant correlation with QoL. [54] Furthermore, by multiple linear regression, miscellaneous symptoms were significantly correlated with QoL in Hispanic patients. [57]

## Discussion

Ethnicity represents a social construct that could be defined as an individual’s sense of culture and Individuals in the same ethnic group often share linguistic, dietary, and religious traits, and potentially share similar outlooks on health and healthcare (Cooper, 1994).. Ethnicity could also be defined as “the fact or state of belonging to a social group that has a common national or cultural tradition”. [59]

Race, ethnicity, and culture have no standard scientific definitions, making these variables difficult to measure and often mixed or used interchangeably. A frequent question arises as to whether meaningful comparative research can be conducted when there is such a considerable likelihood of misclassification [60]

Racial and ethnic categories are frequently used in biomedical research. The importance of application of precise classification remains an area of a large debate along years. The currently used grouping of ethnicities varies in different studies. [61] Ethnicity could be classified according to the population sharing same environmental and social factors as it can be a self-defined or subjective identity. [62] However, with the advancing knowledge of human genetics and variation of gene mapping, the application of currently used ethnic categorizations in medical research become increasingly debatable. [63]

The commonly reported ethnicity classification in medical research is based more on geographical distribution and similarities between certain populations with each ethnicity representing people from many close countries. Among major ethnicities widely represented in different research, Caucasian, with Mediterranean representing subrace of Caucasian, African, Asian, Middle Eastern and Hispanic. [61]

For the first part of the research question, it was clearly reported that NMS affected the QoL in all ethnic categories and exhibited a significant correlation with QoL in all studies included in this review. This confirms the conclusions of previous studies over the past two decades regarding the impact of NMS on QoL, and that NMS is a major determinant of QoL in PD. [64] Moreover, by regression analysis, most of the included studies reported that NMS explained more of the variability in QoL scoring than motor symptoms; therefore, its impact on QoL was greater than the motor symptoms, which is similar to what was reported in literature. Only one study from Turkey found that motor symptoms have a greater impact on QoL than NMS [55]

QoL scores varied among different ethnicities. North African and Middle Eastern groups experienced worse QoL than other ethnicities. Caucasian patients showed a better QoL, whereas East Asian and Mediterranean groups had the best QoL. This indicates that ethnicity might play a role in the way in which PD affects the QoL. However, this variation could be explained by the differences in motor complications experienced by the patients, as well as the NMS impact. Future research on the ethnicity impact on QoL would explain what role and to what extent ethnicity can modify the effect of different PD symptoms on the QoL of PD patients in all parts of the world.

By analysing the correlations between NMS domains and QoL, there was a variation in the impact of certain NMS on QoL between ethnicities. However, some NMS features appeared to impact the QoL significantly in all ethnic categories. Depression was among the most significant NMS features that correlated with QoL in seven out of eight ethnicities. This agrees with what was reported previously by Schrag, (2006b) and other researchers that depression is the principal factor associated with poor HRQoL in PD. Interestingly, depression showed significant but weak correlation with QoL in Iranian patients. Depression also showed a non-significant correlation with QoL in Slovakian patients, However, when depression was assessed in two groups of patients, it correlated significantly with QoL in patients with fluctuating symptoms, while no significant impact was detected in patients without fluctuations. This points out to the need of further research to demonstrate the impact of symptoms fluctuation on the depression.

The impact of sleep and fatigue on QoL was common among all ethnic groups. Similarly, attention and memory had a significant correlation with QoL in all ethnicities except for Mediterranean and South Asian people. This implies that the most frequent NMS, that significantly affect the QoL in PD in most of the ethnicities are mood (particularly depression) followed by sleep/fatigue, and attention /memory problems.

The impact of other non-motor features on QoL was variable between different ethnicities. Urinary symptoms correlated with QoL moderately in North and West African studies, South Asian and one East Asian study[50] [49][45], (Lee *et al.*, 2015)

In addition, Gastrointestinal symptoms correlated with QoL moderately in East Asian, South Asian (one study),[46] Caucasian (English patients only), [47] Middle Eastern North and West African groups. [55], [56] [50] [49]

Interestingly, the only study that reported the sexual dysfunction as one of the major symptoms impacting QoL is from Middle Eastern, in particular, Turkey. [55], This impact was more prominent in PD patients who are younger than 60-year-old. [55],Furthermore, Nigerian and South Asian identified a significant moderate correlation between this domain and QoL. [50] [45]

Middle Eastern, East, and South Asian appeared to be more affected by hallucinations and perceptual problems than other ethnicities. Moreover, cardiovascular symptoms were associated with poor QoL in Mexican, Malaysian[57], [53]. CVS related symptoms were reported as the major NMS impacting the QoL in Mexican and Malaysian study populations. Furthermore, two East Asian and one South Asian studies reported significant correlation with QoL [44] [46] [52]. Miscellaneous symptoms, including pain, loss of taste/smell, weight loss and excessive sweating were correlated with QoL in some patients within the reviewed ethnicities including two studies from East Asia, in addition to Turkish and Mexican patients. Nevertheless, in Slovak patients, pain and DDS were assessed separately, and pain had the most significant impact on QoL, in Slovak patients followed by DDS. [54]

Among the NMSs which reported as determinants of QoL are autonomic dysfunction related symptoms. A study from the UK by [65] reported that autonomic dysfunction was associated with poor QoL in PD. In addition, a well-known Italian study, the PRIAMO, which included a very large sample size concluded that apathy is the NMS causing significant impact on the QoL. [66][67]Furthermore, an international study carried across centres in 10 countries reported that mood/apathy, sleep/fatigue, and miscellaneous domains were the most significant factors contributing to the QoL. [64]

This wide variability in the results reflects that some NMSs affect the QoL in almost all ethnic ethnicities. These symptoms include mood and cognition, sleep, and memory problems. However, there are notable differences in the impact of the other six NMS domains on QoL. The data suggests that ethnicity may play a role in the way certain NMS impact QoL of patients with PD, only in certain ethnic categories.

The prevalence of each NMS was reported in all ethnicities. Some non-motor symptoms were common in all ethnicities, for example, mood and apathy, particularly depression, sleep and fatigue, memory and attention problems, and miscellaneous symptoms’ domain. However, differences in the percentage of prevalence of these symptoms between different ethnicities were observed. In addition, it was reported that Asian PD patients might express mor GIT symptoms than other ethnicities [29] however, only two East Asian studies reported high prevalence of GIT symptoms in their population while the remaining 7 East Asian studies have not reported a very high prevalence in comparison to patients from other ethnicities. Moreover, what is concluded from the impact of certain NMS on QoL comparing to their prevalence, is that the high prevalence of NMS in certain population does not necessitate that it is the most NMS which can significantly impact the QoL. An example of that, in Turkish people with PD, the most 54 common NMS are miscellaneous symptoms domain followed by urinary and mood/cognition domains. However, the two NMS which reported to significantly impact the QoL are sexual dysfunction and perceptual problems.

Some studies have evaluated the correlation between NMS and disease duration, severity, and motor symptoms. NMS were found to be correlated with disease severity and progression assessed, and severity of motor symptoms assessed by UDPRS III in these studies.

In addition to the clinical manifestations that have a major impact on the quality of life, other factors have been evaluated for their impact on QoL, including: disability [20]; patient-related features such as personality traits [68]; psychological adjustment to the disease [69]; and educational, behavioural and psychological factors [70] that are associated with patients’ perceived HRQoL

In this review, we have included certain scales for measuring the QoL and NMS. PDQ39 was selected because it is valid, translated into many languages, and specific to QoL in Parkinson’s disease. For the non -motor symptoms all the reporting scales defined in the inclusion criteria are validated and have worldwide application and are translated into different languages. These include NMSS, NMSQ and MDS -UDPRS I. HADS was also included as a validated instrument for assessment of depression and anxiety, although was only used in one study.

What was clearly noticed during the research of ethnicity definition and classification for the purpose of this review is that there is an inconsistency and a long debate over the most reliable ethnicity categorization. Ethnicity is commonly defined as a social and self-defining grouping, which can be misleading and can result in inaccurate identifying of health behaviours and risks. Thus, trying to define ethnicity is very difficult if we aimed to achieve a good understanding of ethnic variations and precise application of health management accordingly.

## Limitations

- Only one scale was included for the assessment of QoL, the PDQ39. For this reason, we excluded many studies that had not reported the QoL by this scale. This reflects the need for future systematic reviews including more QoL assessment scales to ensure the inclusion of studies from more countries and ethnic groups.
- Although met-regression was the most appropriate analysis for evaluating the impact of non -motor symptoms on the quality of life in this review, it was not possible to perform this type of analysis due to the difficulty in obtaining the raw data of the participants in different studies.
- The heterogeneity in reported scores either as mean±SD or median (IQR), and the differences in scales measuring NMS in different studies, make it difficult to compare all included ethnicities for the differences in both QoL and total NMS scores.
- To this review, the ethnic grouping was based on the country where the study is conducted unless including of patients from different countries was stated by the author.

## Recommendations

The motor symptoms should be assessed concurrently with the NMS to understand the exact relation between the NMS and the motor severity in PD. To avoid any inconsistency and inaccuracy in the assessment of different motor and non -motor symptoms, reporting whether the patients were assessed in On or Off state of their medications is crucial.

In addition, unifying the ethnicity definition and the classification of ethnic groups is essential to avoid the current discrepancies in ethnic categorization. The precise reporting of ethnicity in biomedical research is crucial for the relevant understanding of many diseases in different ethnicities and application of appropriate management plans. Future studies should consider ethnicity when reporting the results of the included populations. This would facilitate our achievement of more understanding of the relation between ethnicity and PD in all areas. Such areas include motor and non-motor symptoms, motor complications, response to treatment, staging, impact of age, disease duration and other demographics on the disease progression and QoL in different ethnicities.

## Conclusion

We demonstrated in this review that there are convincing evidence of ethnic variabilities in NMS and QoL, in addition ethnicity can be a predictor of NMS and QoL in patients with PD. PD patients from North African and middle eastern origin appear to experience the highest NMS and the worst QoL, while East Asian and Mediterranean patients are experiencing less burden of NMS and a better QoL. Depression, sleep difficulties, fatigue, attention, and memory problems are the commonest NMS features that have a major impact on QoL in most ethnicities. No studies assessed the effect of NMS in QoL of Arab, South African, South American (except Brazil). It worth noting, this review did not address how and why the ethnicity impact PD symptoms. Further research in this area including more large multicentre quantitative and qualitative studies including patients from different ethnicities are needed to air our understanding of the ethnic variabilities in PD symptoms and their effect in QoL.

## Data Availability

All data produced in the present work are contained in the manuscript

