## Appendix 1 & 2 for "Ethnicity as a predictor of nonmotor symptoms impact on quality of life in patients with Parkinson’s disease: A systematic review"

Appendix 1 Summary of database search strategy *(last reviewed 15th September 2023)*

| **Syntax used for NMS and QoL in PD search** |
|---|
| **PubMed search (Search 1)** |
| **("Parkinson disease"[MeSH Terms] OR "Parkinson"[tw] OR "Parkinson's"[tw] OR "Parkinsons"[tw]) AND ("quality of life"[MeSH Terms] OR "quality of life") AND ("questionnaire" OR "questionnaires" OR "rating" OR "PDQ-39" OR "HADS-P" OR "UPDRS" OR "NMSS")** |
| **Search yielded (1808 ) results** |
| **Ovid search (search 2)** |
| **1 exp Parkinson disease/ or (Parkinson or Parkinson s or Parkinson's).ti,ab.**<br>**2 exp "quality of life"/ or quality of life.ti,ab.**<br>**3 (questionnaire or questionnaires or rating or PDQ-39 or HADS-P or UPDRS or NMSS).ti,ab.**<br>**4 1 and 2 and 3** |
| **Search yielded (4264 )results** |
| **Scopus search (search 3)** |
| **( TITLE-ABS-KEY ( parkinson OR parkinsons OR parkinson's ) AND TITLE-ABS-KEY ( "quality of life" ) AND TITLE-ABS-KEY ( questionnaire OR questionnaires OR rating OR "PDQ-39" OR "HADS-P" OR "UPDRS" OR "NMSS" )** |
| **Search yielded ( 2910 ) results** |
| **Total search results (8982)** |

### APPENDIX 2 QUALITY ASSESSMENT OF INCLUDED STUDIES.

| No. | Criteria | Maximum score | (Liu et al., 2015) | (Duncan et al., 2014) | (Rodriguez-Violante et al., 2013) | (Salar et al., 2017) | (Berganzo et al., 2016) | (Shalash et al., 2018) | (Kwon et al., 2016) | (Arabambi, Oshinaike and Ogun, 2019) | (Prakash, 2016) | (Li et al., 2010) | (Song et al., 2014) | (Wu et al., 2016) | (Azmin et al., 2014) | (Hu et al., 2018) | (Lee et al., 2015) | (L Kadastik-Eerme et al., 2016)Eerme, 2015) | Skorvanek, 2015) | (Pappala et al., 2019) | (Gulunay et al., 2020) | (Karri, Ramasamy and Kalidoss, 2020) |
|---|---|---|---|---|---|---|---|---|---|---|---|---|---|---|---|---|---|---|---|---|---|---|
| 1 | **Study objectives** | 1 | 1 | 1 | 1 | 1 | 1 | 1 | 1 | 1 | 1 | 1 | 1 | 1 | 1 | 1 | 1 | 1 | 1 | 1 | 1 | 1 |
| 2 | **Study design** | 1 | 1 | 1 | 1 | 1 | 1 | 1 | 1 | 1 | 1 | 1 | 1 | 1 | 1 | 1 | 1 | 1 | 1 | 1 | 1 | 1 |
| 3 | **Cognition of studied population** | 2 | 0 | 1 | 0 | 1 | 0 | 1 | 1 | 0 | 1 | 2 | 2 | 2 | 1 | 2 | 2 | 2 | 1 | 0 | 2 | 2 |
| 4 | **Characteristics of studied ppopulation** | 2 | 1 | 2 | 2 | 2 | 2 | 2 | 2 | 1 | 2 | 2 | 2 | 2 | 1 | 2 | 2 | 2 | 2 | 1 | 2 | 2 |
| 5 | **Sampling method for recruitment of study population** | 2 | 2 | 2 | 2 | 2 | 2 | 2 | 2 | 2 | 2 | 2 | 2 | 2 | 2 | 2 | 2 | 2 | 2 | 2 | 2 | 2 |
| 6 | **Sample size** | 2 | 2 | 1 | 1 | 0 | 1 | 0 | 2 | 1 | 2 | 0 | 2 | 2 | 1 | 2 | 1 | 2 | 2 | 0 | 0 | 1 |
| 7 | **Definition of the concept of HRQoL** | 1 | 0 | 0 | 0 | 0 | 0 | 0 | 0 | 0 | 0 | 1 | 1 | 0 | 0 | 0 | 0 | 0 | 0 | 0 | 0 | 0 |
| 8 | **Justified choice of HRQoL-assessing instrument** | 2 | 2 | 2 | 2 | 2 | 1 | 1 | 1 | 2 | 2 | 1 | 2 | 2 | 2 | 1 | 1 | 2 | 2 | 2 | 2 | 2 |
| 9 | **Justified choice of NMS-assessing instrument** | 2 | 2 | 2 | 2 | 2 | 1 | 1 | 1 | 2 | 2 | 1 | 2 | 2 | 2 | 1 | 1 | 2 | 2 | 2 | 2 | 2 |
| 10 | **Comprehensible statistical methods** | 1 | 1 | 1 | 1 | 1 | 1 | 1 | 1 | 1 | 1 | 1 | 1 | 1 | 1 | 1 | 1 | 1 | 1 | 1 | 1 | 1 |
| 11 | **Main factors associated with HRQoL** | 1 | 1 | 1 | 1 | 1 | 1 | 1 | 1 | 1 | 1 | 1 | 1 | 1 | 1 | 1 | 1 | 1 | 1 | 1 | 1 | 1 |
| 12 | **Main NMS features associated with HRQoL** | 1 | 1 | 1 | 1 | 0 | 1 | 1 | 1 | 1 | 1 | 1 | 1 | 1 | 1 | 1 | 1 | 1 | 1 | 0 | 1 | 1 |
| 13 | **Agreement / disagreement with other studies** | 2 | 2 | 2 | 2 | 2 | 2 | 2 | 2 | 2 | 1 | 2 | 2 | 2 | 2 | 2 | 2 | 2 | 2 | 2 | 2 | 2 |
| 14 | **Limitations** | 1 | 1 | 1 | 1 | 1 | 1 | 1 | 1 | 1 | 1 | 1 | 0 | 1 | 1 | 1 | 1 | 1 | 1 | 1 | 1 | 1 |
| **Total score** | | 21 | 17 | 18 | 17 | 17 | 15 | 15 | 17 | 16 | 18 | 17 | 20 | 20 | 17 | 18 | 17 | 20 | 19 | 14 | 18 | 19 |
| **Comment** | | Good | Good | Good | Good | Good | Good | Good | Good | Good | [illegible] | [illegible] | [illegible] | [illegible] | Good | [illegible] | Good | Good | [illegible] | Fair | Good | Good |

| Ethnicity | Study | CVS | Sleep/ Fatigue | Mood/ Cognition | Perception | Attention Memory | GIT | Urinary | Sexual function | Other |
|---|---|---|---|---|---|---|---|---|---|---|
| East Asian | (Li et al., 2010) | 0.109 | 0.34 | 0.69 | 0.212 | 0.202 | 0.384 | 0.248 | 0.006 | 0.323 |
| | (Song et al., 2014) | 0.28 | 0.6 | 0.55 | 0.3 | 0.42 | 0.44 | 0.33 | 0.12 | 0.46 |
| | (Wu et al., 2016) | 0.258 | 0.561 | 0.659 | 0.19 | 0.46 | 0.333 | 0.119 | 0.075 | 0.321 |
| | (Hu et al., 2018) | 0.206 | 0.527 | 0.573 | 0.19 | 0.333 | 0.277 | 0.193 | 0.078 | 0.298 |
| | (Kwon et al., 2016) | 0.457 | 0.528 | 0.573 | 0.343 | 0.425 | 0.43 | 0.363 | 0.26 | |
| | (Lee et al., 2015) | 0.305 | 0.559 | 0.669 | 0.313 | 0.557 | 0.383 | 0.435 | 0.335 | 0.735 |
| | (Liu et al., 2015) | 0.403 | 0.461 | 0.607 | 0.371 | 0.411 | 0.383 | 0.172 | 0.253 | 0.258 |
| | (Prakash, 2016) | | 0.30* | 0.32* | | 0.63* | | | | |
| | (Azmin et al., 2014) | 0.563 | 0.469 | 0.588 | 0.336 | 0.281 | 0.412 | 0.154 | 0.173 | 0.258 |
| South Asian | (Pappala et al., 2019) | 0.24 | 0.54 | 0.49 | 0.49 | 0.28 | 0.35 | 0.25 | 0.4 | 0.26 |
| Caucasian | (Duncan et al., 2014) | | 0.438 | 0.307** | | 0.2** | 0.15** | | | |
| | (Skorvanek, 2015) | | 0.18** | | | | | | | 0.25** |
| Mediterranean | (Berganzo et al., 2016) | | 0.44 | 0.54 | | 0.24 | | 0.24 | 0.24 | 0.17 |
| Hispanic | (Rodriguez-Violante et al., 2013) | | 0.51 | 0.52 | | 0.53 | | | | |
| Middle Eastern | (Gulunay et al., 2020) | | | 0.66 | 0.72 | | 0.247 | | 0.472 | 0.232 |
| | (Salar et al., 2017) | 0.11 | 0.363 | 0.377 | 0.392 | 0.395 | 0.11 | 0.158 | 0.12 | 0.116 |
| North African | (Shalash et al., 2018) | 0.198 | 0.659 | 0.681 | 0.263 | 0.421 | 0.452 | 0.414 | 0.151 | - |
| West African | (Arabambi, Oshinaike and Ogun, 2019) | 0.556 | 0.713 | 0.74 | 0.417 | 0.353 | 0.533 | 0.621 | 0.494 | 0.539 |
